# De novo and inherited dominant variants in U4 and U6 snRNAs cause retinitis pigmentosa

**DOI:** 10.1101/2025.01.06.24317169

**Authors:** Mathieu Quinodoz, Kim Rodenburg, Zuzana Cvackova, Karolina Kaminska, Suzanne E de Bruijn, Ana Belén Iglesias-Romero, Erica G M Boonen, Mukhtar Ullah, Nick Zomer, Marc Folcher, Jacques Bijon, Lara K Holtes, Stephen H Tsang, Zelia Corradi, K Bailey Freund, Stefanida Shliaga, Daan M Panneman, Rebekkah J Hitti-Malin, Manir Ali, Ala’a AlTalbishi, Sten Andréasson, Georg Ansari, Gavin Arno, Galuh D N Astuti, Carmen Ayuso, Radha Ayyagari, Sandro Banfi, Eyal Banin, Mirella T S Barboni, Miriam Bauwens, Tamar Ben-Yosef, David G Birch, Pooja Biswas, Fiona Blanco-Kelly, Beatrice Bocquet, Camiel J F Boon, Kari Branham, Alexis Ceecee Britten-Jones, Kinga M Bujakowska, Elizabeth L Cadena, Giacomo Calzetti, Francesca Cancellieri, Luca Cattaneo, Peter Charbel Issa, Naomi Chadderton, Luísa Coutinho-Santos, Stephen P Daiger, Elfride De Baere, Berta de la Cerda, John N De Roach, Julie De Zaeytijd, Ronny Derks, Claire-Marie Dhaenens, Lubica Dudakova, Jacque L Duncan, G Jane Farrar, Nicolas Feltgen, Lidia Fernández-Caballero, Juliana M Ferraz Sallum, Simone Gana, Alejandro Garanto, Jessica C Gardner, Christian Gilissen, Kensuke Goto, Roser Gonzàlez-Duarte, Sam Griffiths-Jones, Tobias B Haack, Lonneke Haer-Wigman, Alison J Hardcastle, Takaaki Hayashi, Elise Héon, Alexander Hoischen, Josephine P Holtan, Carel B Hoyng, Manuel Benjamin B Ibanez, Chris F Inglehearn, Takeshi Iwata, Kaylie Jones, Vasiliki Kalatzis, Smaragda Kamakari, Marianthi Karali, Ulrich Kellner, Krisztina Knézy, Caroline C W Klaver, Robert K Koenekoop, Susanne Kohl, Taro Kominami, Laura Kühlewein, Tina M Lamey, Bart P Leroy, María Pilar Martín-Gutiérrez, Nelson Martins, Laura Mauring, Rina Leibu, Siying Lin, Petra Liskova, Irma Lopez, Victor R de J López-Rodríguez, Omar A Mahroo, Gaël Manes, Martin McKibbin, Terri L McLaren, Isabelle Meunier, Michel Michaelides, José M Millán, Kei Mizobuchi, Rajarshi Mukherjee, Zoltán Zsolt Nagy, Kornelia Neveling, Monika Ołdak, Michiel Oorsprong, Yang Pan, Anastasia Papachristou, Antonio Percesepe, Maximilian Pfau, Eric A Pierce, Emily Place, Raj Ramesar, Florence Andrée Rasquin, Gillian I Rice, Lisa Roberts, María Rodríguez-Hidalgo, Javier Ruiz-Eddera, Ataf H Sabir, Ai Fujita Sajiki, Ana Isabel Sánchez-Barbero, Asodu Sandeep Sarma, Riccardo Sangermano, Cristina M Santos, Margherita Scarpato, Hendrik P N Scholl, Dror Sharon, Sabrina Giovanna Signorini, Francesca Simonelli, Ana Berta Sousa, Maria Stefaniotou, Katarina Stingl, Akiko Suga, Lori S Sullivan, Viktória Szabó, Jacek P Szaflik, Gita Taurina, Carmel Toomes, Viet H Tran, Miltiadis K Tsilimbaris, Pavlina Tsoka, Veronika Vaclavik, Marie Vajter, Sandra Valeina, Enza Maria Valente, Casey Valentine, Rebeca Valero, Joseph van Aerschot, L. Ingeborgh van den Born, Andrew R Webster, Laura Whelan, Bernd Wissinger, Georgia G Yioti, Kazutoshi Yoshitake, Juan C Zenteno, Roberta Zeuli, Theresia Zuleger, Chaim Landau, Allan I Jacob, Frans P M Cremers, Winston Lee, Jamie M Ellingford, David Stanek, Carlo Rivolta, Susanne Roosing

## Abstract

The U4 small nuclear RNA (snRNA) forms a duplex with the U6 snRNA and, together with U5 and ∼30 proteins, is part of the U4/U6.U5 tri-snRNP complex, located at the core of the major spliceosome. Recently, recurrent *de novo* variants in the U4 RNA, transcribed from the *RNU4-2* gene, and in at least two other *RNU* genes were discovered to cause neurodevelopmental disorder. We detected inherited and *de novo* heterozygous variants in *RNU4-2* (n.18_19insA and n.56T>C) and in four out of the five *RNU6* paralogues (n.55_56insG and n.56_57insG) in 135 individuals from 62 families with non-syndromic retinitis pigmentosa (RP), a rare form of hereditary blindness. We show that these variants are recurrent among RP families and invariably cluster in close proximity within the three-way junction (between stem-I, the 5’ stem-loop and stem-II) of the U4/U6 duplex, affecting its natural conformation. Interestingly, this region binds to numerous splicing factors of the tri-snRNP complex including PRPF3, PRPF8 and PRPF31, previously associated with RP as well. The U4 and U6 variants identified seem to affect snRNP biogenesis, namely the U4/U6 di-snRNP, which is an assembly intermediate of the tri-snRNP. Based on the number of positive cases observed, deleterious variants in *RNU4-2* and in *RNU6* paralogues could be a significant cause of isolated or dominant RP, accounting for up to 1.2% of all undiagnosed RP cases. This study highlights the role of non-coding genes in rare Mendelian disorders and uncovers pleiotropy in *RNU4-2*, where different variants underlie neurodevelopmental disorder and RP.

## MAIN TEXT

While approximately 2 million individuals are affected by retinitis pigmentosa (RP) worldwide, it is estimated that 30-50% of these individuals remain without a conclusive genetic diagnosis, even when exome and/or genome sequencing is performed^1-4^. This is due, in part, to the high genetic heterogeneity of the disease, limited opportunities for genetic testing, and the existence of yet-to-be-identified causal genes. Moreover, the identification of novel disease-associated genes is hindered because RP variants are exceedingly rare in the population, with 97% and 81% occurring at allele frequencies below 0.1% and 0.01%, respectively, in any control subpopulations^5,6^.

Non-coding RNAs are essential to many regulatory cellular processes, such as the regulation of gene expression and pre-mRNA gene splicing^7^. In particular, five small nuclear RNAs (snRNAs), U1, U2, U4, U5, and U6, are constituents of the major spliceosome, a large and dynamic macromolecular complex involved in pre-mRNA splicing, a critical intermediate step between transcription and translation for the vast majority of genes in the human genome. Recently, recurrent *de novo* variants in *RNU4-2*, one of the two paralogues encoding the human U4 snRNA, have been associated with a frequent neurodevelopmental disorder (NDD), ReNU syndrome (OMIM: 620851). Pathogenic variants in this gene have been shown to cause systematic bias in the recognition of the donor splice site by the spliceosome and to account for as much as 0.4% of all individuals with NDD^8,9^. Likewise, other snRNA genes, *RNU2-2P* and *RNU5B-1*, have also been recently described to be linked to the same condiiton^10-12^.

Several spliceosomal proteins are also known to be involved in a wide range of hereditary diseases, including RP. Specifically, out of the 114 genes (https://retnet.org/) that are currently associated with non-syndromic RP, the tri-snRNP splicing factors genes *PRPF3, PRPF4, PRPF6, PRPF8, PRPF31*, and *SNRNP200* underly the autosomal dominant form of the condition (adRP), with variants in *PRPF31* accounting for 10-20% of all such cases^3,13^.

Here, we identified both inherited and *de novo* variants in *RNU4-2* and four paralogues of *RNU6*, encoding the U6 snRNA, as the molecular cause of adRP in 135 individuals across 62 families. We demonstrate that all identified variants cluster in close proximity, within the U4/U6 duplex, and specifically in a region that binds directly to PRPF31 and PRPF3, and indirectly to PRPF6 and PRPF8^14,15^. Finally, we show that the RP-associated variants increase the association of U4 and U6 snRNAs with di-snRNP-specific proteins SART3 and PRPF31.

### *RNU4-2* variants underlie adRP

We initially examined a non-consanguineous family with adRP spanning two generations. Seven of eight siblings (II:1-II:7) and their father (I:1) were affected, each experiencing a symptomatic onset of night-blindness and progressive loss of peripheral vision beginning in late adolescence to early adulthood. Fundus examination and electrophysiological testing of all affected individuals revealed classical RP features (Supplementary Fig. 1, Supplementary data 1). Genome sequencing was performed for all ten individuals of the family and selective DNA variant filtering and shared haplotype analysis revealed the presence of a variant in *RNU4-2* (NR_003137.2:n.18_19insA, Supplementary Table 1) as the possible cause of disease. Among other features, this variant was absent from genomic databases of controls and was present in a highly conserved region of the genome^6,16-18^.

We then extended our analysis to 1,893 individuals with RP without prior genetic diagnosis and identified three additional families, consisting of 15 affected individuals segregating the same pathogenic variant. In addition, we detected another DNA variant, n.56T>C, that recurred in seven individuals from four families (Supplementary Table 1 and 2, Fig. 1A,C). Similar to n.18_19insA, the n.56T>C variant was absent from genomic databases of control populations. Additional screening of patient cohorts from our respective institutions, as well as the analysis of the UK National Genomic Research Library, hosting data from the Genomics England 100,000 Genomes Project (GEL)^19^, and from the NHS Genomic Medicine Service (GMS), comprising a total of ∼3,000 cases, uncovered an additional patient harboring n.18_19insA and four families (five affected individuals) carrying the n.56T>C variant. Altogether, recurrent variants in *RNU4-2* were identified in 36 affected individuals from 13 families (Supplementary Fig. 2). Twenty-seven additional affected individuals from these families could not be assessed genetically as no DNA was available. Of note, incomplete penetrance was observed for nine obligate carriers (of which four were deceased) in 38% of families (five out of 13), who would bear pathogenic variants but have no visual symptoms (Supplementary Fig. 1). One of these individuals was shown to be subclinically affected, one showed no clinical signs of disease upon clinical examination, and three were not clinically evaluated to confirm their unaffected status.

**Figure 1:**
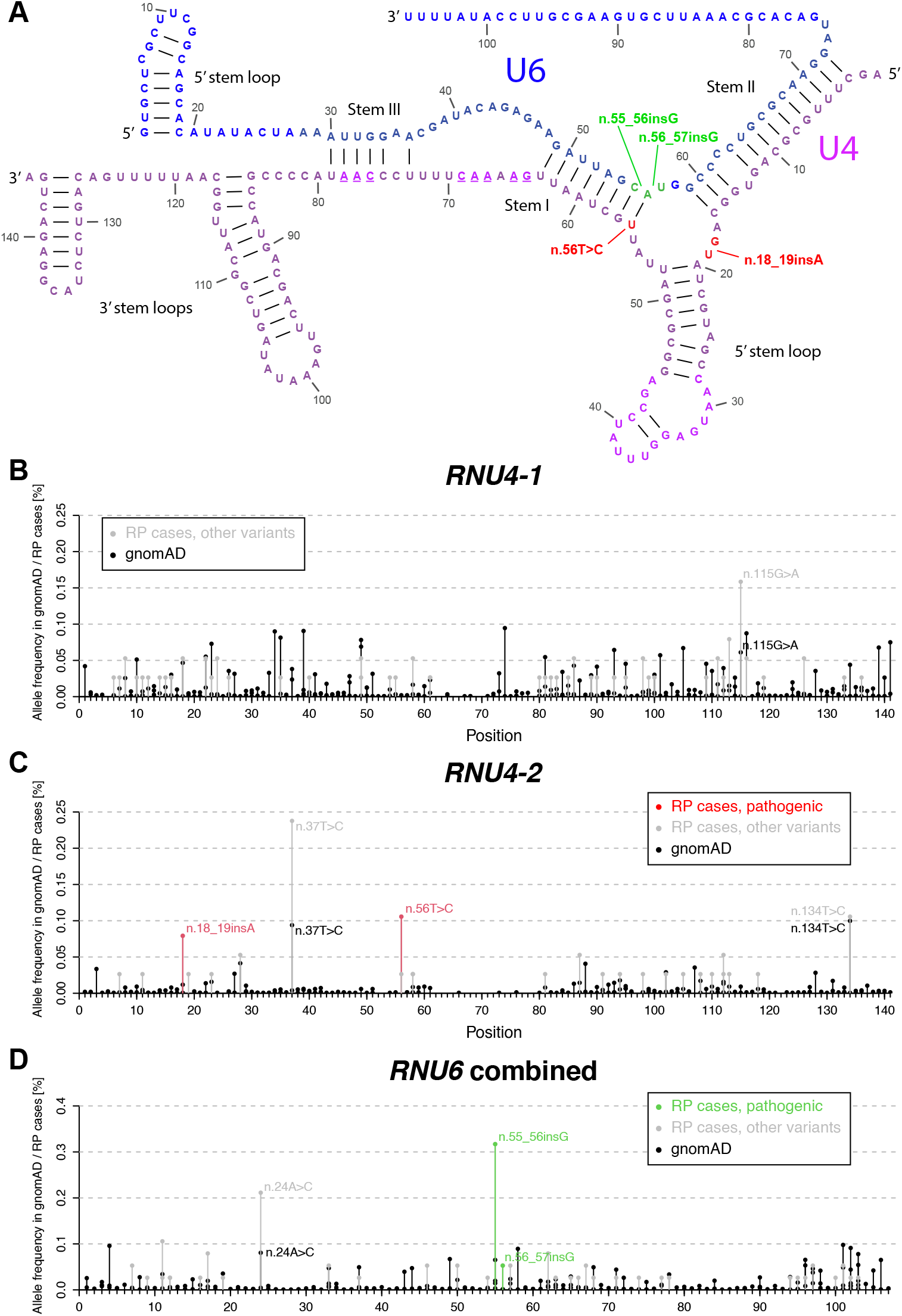
U4-U6 structure and rare variants found in RP cases and controls (gnomAD). (A) 2D structure of U4-U6 duplex with recurrent variants identified in RP cases (in red for U4 and in green for U6), which cluster in the same region of the complex, the three-way junction. Nucleotides affected by variants previously observed in neurodevelopmental disorder cases are underlined. (B) Rare variants affecting *RNU4-1*, defined as AF<0.1% in gnomAD v.4.1, identified in RP cases and in controls. (C) same as (B) for *RNU4-2* with recurrent variants displayed in red. (D) same as (B) for all five *RNU6* paralogues combined, with recurrent variants displayed in green.

Since the only U4 paralogue, *RNU4-1*, differs from *RNU4-2* only at four positions (n.37, n.88, n.99, and n.113), we also assessed this gene (Supplementary Table 3). We did not identify any potentially pathogenic variants, including variants at the sites corresponding to n.18_19 and n.56 of *RNU4-2* (Fig. 1B). Conversely, 63 likely benign variants were detected (Supplementary Table 4). NR_003925.1:n.56T>C was observed in two individuals from the “All of Us” database^18^ but we were unable to assess their phenotype. Notably, *RNU4-1* appears to be more tolerant to variation compared to *RNU4-2*, as evidenced by the many and frequent variants that are present in genomes from the general population (cumulative allele frequency of 20.4% in *RNU4-1* vs 1.2% in *RNU4-2*, gnomAD v4.1) (Fig. 1B,C, Supplementary Fig. 3), although these seem to be reduced in the region corresponding to where pathogenic NDD variants were described in *RNU4-2* (Supplementary Fig. 3), indicating that *RNU4-1*-derived U4 could be partly functional.

In addition to n.18_19insA and n.56T>C, we identified 10 other unique rare variants in *RNU4-2* in 10 RP families, which were classified as variants of uncertain significance (VUS), as well as 17 benign changes (Supplementary Table 4).

### Variants in U6 paralogues also cause RP

In the U4/U6.U5 tri-snRNP complex, U4 binds to U6, to form the U4/U6 RNA duplex. Since the pathogenic variants observed in U4 were predicted to affect this RNA-RNA structure, we reasoned that variants in U6 could also be associated with adRP and extended our analysis to all five identical paralogous genes producing U6 RNA, scattered across the genome (*RNU6-1, RNU6-2, RNU6-7, RNU6-8* and *RNU6-9;* Supplementary Table 3). In the same international cohorts of patients that were previously analyzed, we identified variants at two specific sites in U6 (n.55_56insG and n.56_57insG) in all *RNU6* paralogues, except *RNU6-7*. Importantly, these variants were either absent or extremely rare in controls and were identified only in individuals with RP (Fig. 1A,D, Supplementary Fig. 2).

In total, pathogenic and likely pathogenic U6 variants were detected in 99 affected individuals from 49 families; 112 additional affected individuals from the same pedigrees could not be tested, but are likely to carry the same pathological genotypes. Variant n.55_56insG was present in the majority of the cases (89 individuals from 43 families), occurring in four out of the five *RNU6* paralogues: *RNU6-1, RNU6-2, RNU6-8*, and *RNU6-9* (Supplementary Table 1 and 2, Supplementary Fig. 2). Notably, this variant was confirmed to be a *de novo* event in seven individuals, clinically identified as sporadic cases. In 12 additional pedigrees, n.55_56insG was observed in individuals born to unaffected parents, for which *de novo*-ness was suspected but could not be proven, due to lack of parental DNA.

The nearby U6 variant, n.56_57insG, was identified in *RNU6-1, RNU6-2*, and *RNU6-9*, in six families with adRP (10 individuals, Fig. 1A,D, Supplementary Fig. 2). This variant is observed once in *RNU6-2* in gnomAD v4.1, in a non-Finnish European individual in the age range of 50-55 with an unknown health-status.

Similar to the screening of the *RNU4* paralogues, our analysis of *RNU6* paralogues revealed 20 VUSs and 70 benign variants, validated by Sanger sequencing (Supplementary Table 4).

In summary, we identified variants in *RNU4-2* or *RNU6* paralogues to underlie *de novo* or inherited dominant RP in 62 families. The overall phenotype in all cases in the study cohort was consistent with classical RP on examination and electrophysiological testing with symptomatic onset beginning predominantly in adolescence (Supplementary Table 5). In addition, other concurrent ocular disease features were noted across individuals in the cohort: cystoid macular edema (CME) (55.9%), non-age-related lens opacities (23.6%), and various vitreomacular complications (30.6%) (Supplementary Table 5). Based on our data from ∼5,000 RP cases (most lacking a genetic diagnosis) and taking into consideration that on average ∼40% of people with RP remain genetically undiagnosed, we can estimate that *RNU4* and *RNU6*-associated RP could be responsible for 1.2% of all undiagnosed cases with this disease. Furthermore, considering that ∼30% of RP cases are adRP^20,21^ and 40 adRP families had recurrent variants in *RNU4-2* or *RNU6* paralogues, we can further infer that those variants can account for ∼2.7% of adRP families.

### Predicted effects of variants on the U4/U6 duplex

Analysis of the position of the detected variants in the three-dimensional space of the tri-snRNP complex showed that all variants clustered in close proximity to each other, and in particular within the three-way junction between stem-I and stem-II of the U4/U6 duplex and the 5’ stem-loop of U4 (Fig. 1A). The duplex is stabilized by intramolecular motifs and through base-pairing between U4 and U6 within stem-I (n.56-61), stem-II (n.1-16), stem-III (n.73-79) and the U4 quasi-pseudoknot (n.62 and n.64).

The U4 and U6 variants associated with RP cluster in a different region compared to those underlying NDD (Fig. 1A). *In silico* modeling predicts that the *RNU4-2* variant n.18_19insA inserts a nucleotide between stem-II and the U4 5’ stem-loop affecting the normal base-pairing (Fig. 2A,B), while n.56T>C disrupts the base-pairing between stem-I of the U4/U6 duplex (Fig. 2A,C). Both alterations lead to the extension of the internal loop, an event that was predicted to impact the overall stability of the duplex. In addition, n.18_19insA also modifies the orientation of the 5’ stem-loop relative to stem I and stem II (Fig. 2B).

**Figure 2:**
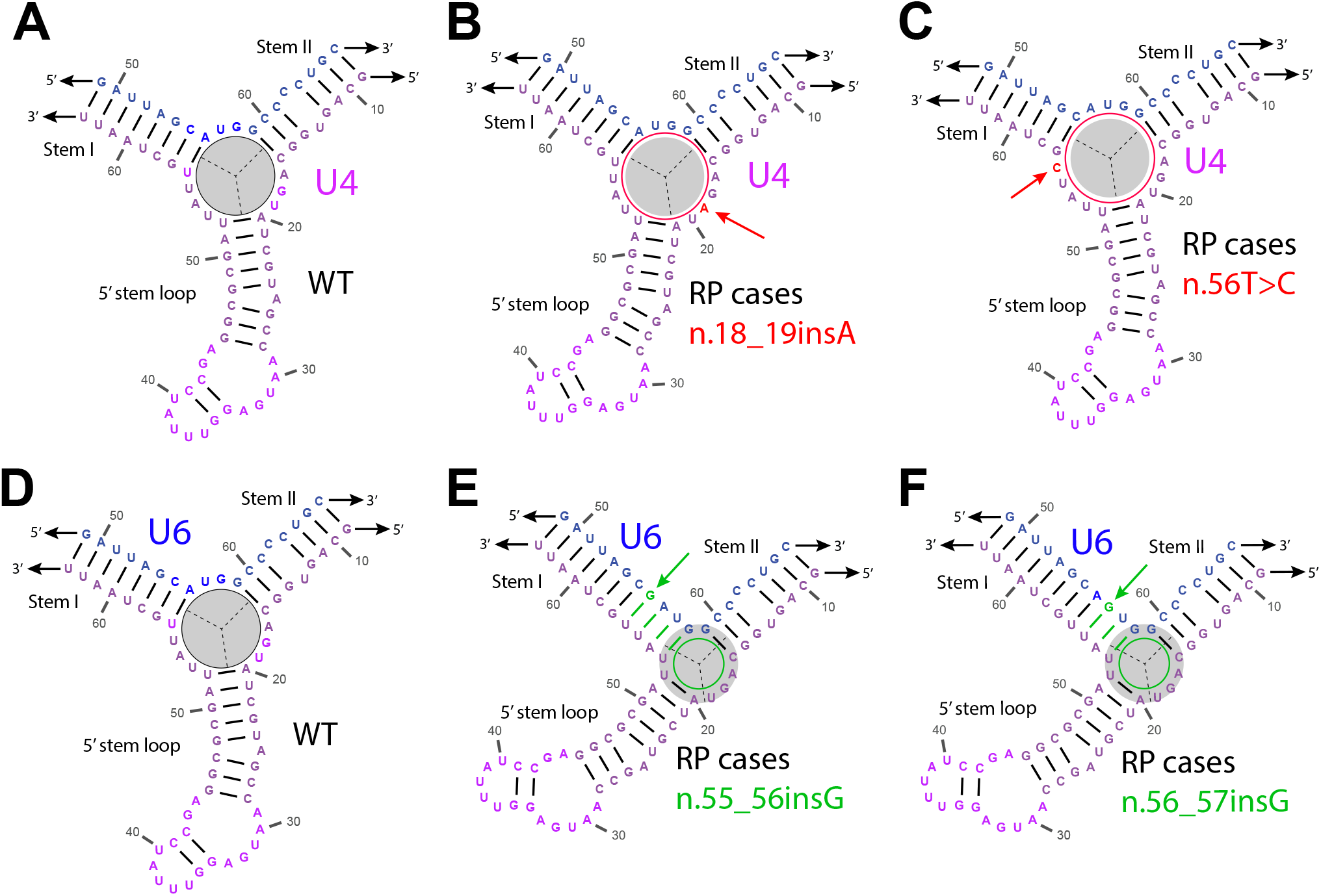
2D modeling of the U4-U6 three-way junction. Wild-type (WT) structure of the U4/U6 duplex surrounding the internal loop (A, D) and structures including pathogenic variants affecting U4 (B, C), U6 (E, F). The gray circle gauges the normal size of the three-way junction, while the dashed lines show the normal orientations of the stems originating from the junction.

In contrast, variants n.55_56insG and n.56_57insG in the *RNU6* paralogs are predicted to introduce an additional wobble base-pairing (G-U) with n.56T of U4 and three additional pairings in stem-I of the U4/U6 duplex. These changes alter the duplex’s secondary structure by extending the length of stem-I, reducing the size of the internal loop, and drastically changing the orientation of the 5’ stem-loop (Fig. 2D-F). Interestingly, we observed that a benign insertion at the same position, n.55_56insT, was present in gnomAD v4.1 in all five *RNU6* paralogues with a cumulative frequency of 0.12% (n=181). Similarly, a nearby variant, n.57T>G was observed in multiple paralogues, in genotypes from the unaffected population (n=21, gnomAD v4.1).

The recurring variants cluster in the three-way junction close to the 5’ stem-loop (Fig. 1A, 3A), which is known to be critical for the binding of the U4/U6 duplex with the splicing factors PRPF31, PRPF3, PRPF6, and PRPF8, all previously associated with adRP (Fig. 3B)^14,15^. Specifically, this structure first binds either PRPF31 or PRPF3/4, and is then stabilized to its original orientation when all factors are bound^22^. The mutated and neighboring U4 and U6 nucleotides in RP cases detected in our study directly participate in the binding of PRPF31 and PRPF3 (Fig. 3C,D), via hydrogen bonds with eight and three residues of these proteins, respectively. Notably, by querying the ClinVar database^23^, we identified a missense variant affecting one of these residues, p.(Arg449Gly) of PRPF3, that was described in a three-generation family of seven affected individuals with an adRP phenotype similar to most of the affected cases in our study cohort^24^.

**Figure 3:**
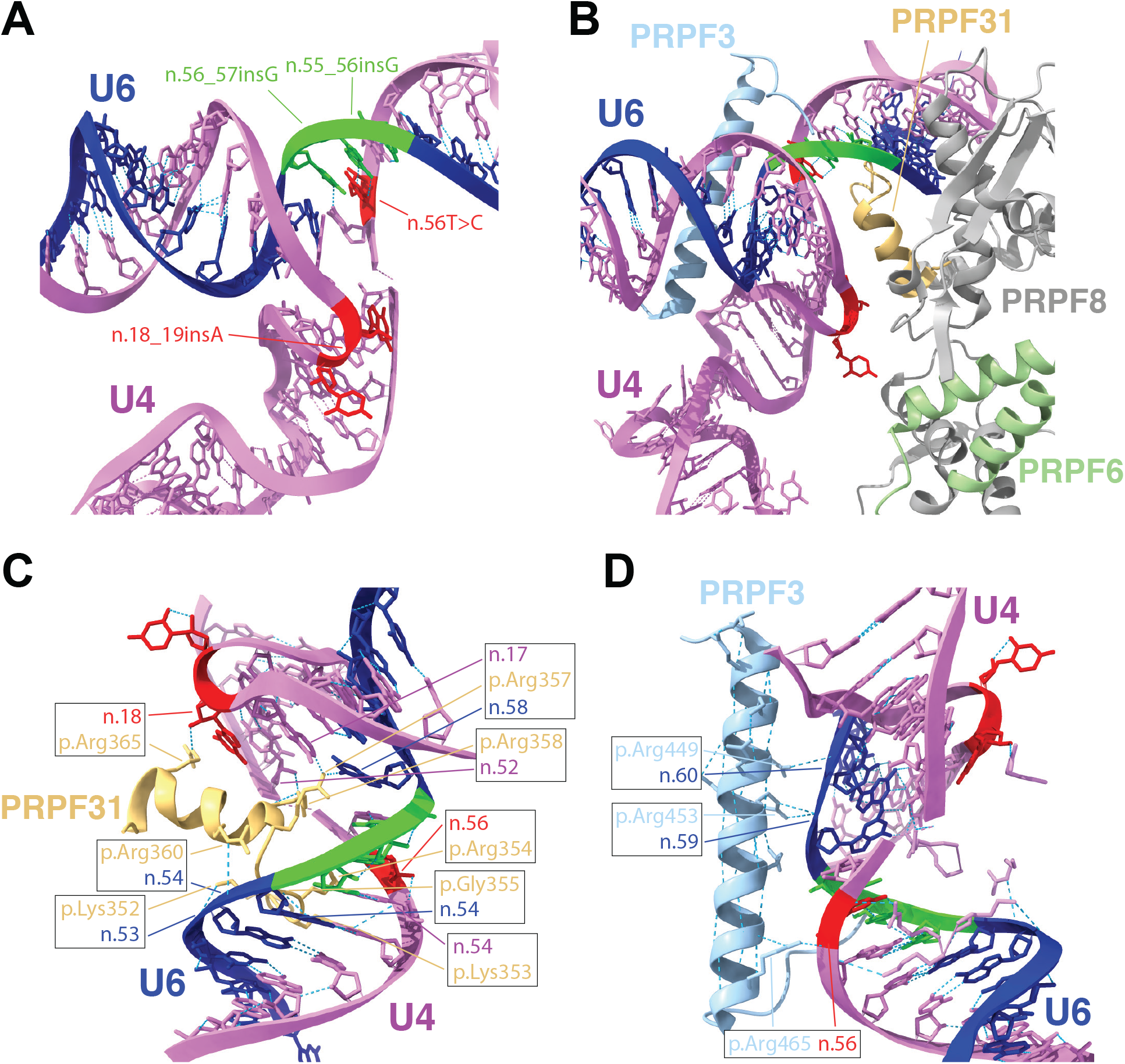
3D structure of the U4/U6 duplex and its interactions with the neighboring splicing factors PRPF3, PRPF6, PRPF8 and PRPF31. (A) Naked U4/U6 pairing, showing the close proximity of the pathogenic variants identified (red and green). (B) Same as in (A), with interacting PRPF proteins. (C) Direct interactions of nucleotides of the U4/U6 duplex with PRPF31, by direct hydrogen bonds. (D) Same as (C) but for PRPF3.

### *RNU4* and *RNU6* genes and their expression

Since the human genome contains several *RNU4* and *RNU6* pseudogenes^25^, we investigated whether any of these might be incorrectly annotated and could instead produce functional RNA, potentially contributing to the disease. In addition, we sought to understand why the various U4 and U6 paralogues appear to be differentially mutated, with *RNU4-1* and *RNU6-7* displaying none of the recurrent pathogenic variants in RP cases. We used RNA-seq data from neurosensory retina (NSR), retinal pigment epithelium (RPE), and human choroid, that had been enriched for small RNA, with bioinformatics analysis performed in a strict manner to mitigate complexities associated with reads aligning against multiple paralogues and/or pseudogenes (see Material and Methods). This analysis showed that *RNU4-2* has a higher expression compared to *RNU4-1* in all tissues analyzed, with an average ratio of 1.65 (Fig. 4A). Conversely, for *RNU6* genes and pseudogenes, individual expression in the retina could not be reliably quantified by RNA-seq, since their sequences are identical, except for the last nucleotide. Therefore, we compared total expression of *RNU4* and *RNU6*, regardless of their respective paralogues and pseudogenes. *RNU6* expression was on average 2.89x higher across the three tissues, compared to *RNU4* (Fig. 4B). Of note, NSR and RPE had higher expression of *RNU4* (2.45x) and of *RNU6* (6.24x) with respect to the choroid, an ocular tissue that is not directly involved in vision and was therefore used as a control (Fig. 4B). This observation is in agreement with previous data, showing that snRNA expression in the retina is approximately 6-fold higher compared to muscle, testis, heart, and brain^26^, indicating a high demand for snRNAs in these two retinal layers.

**Figure 4:**
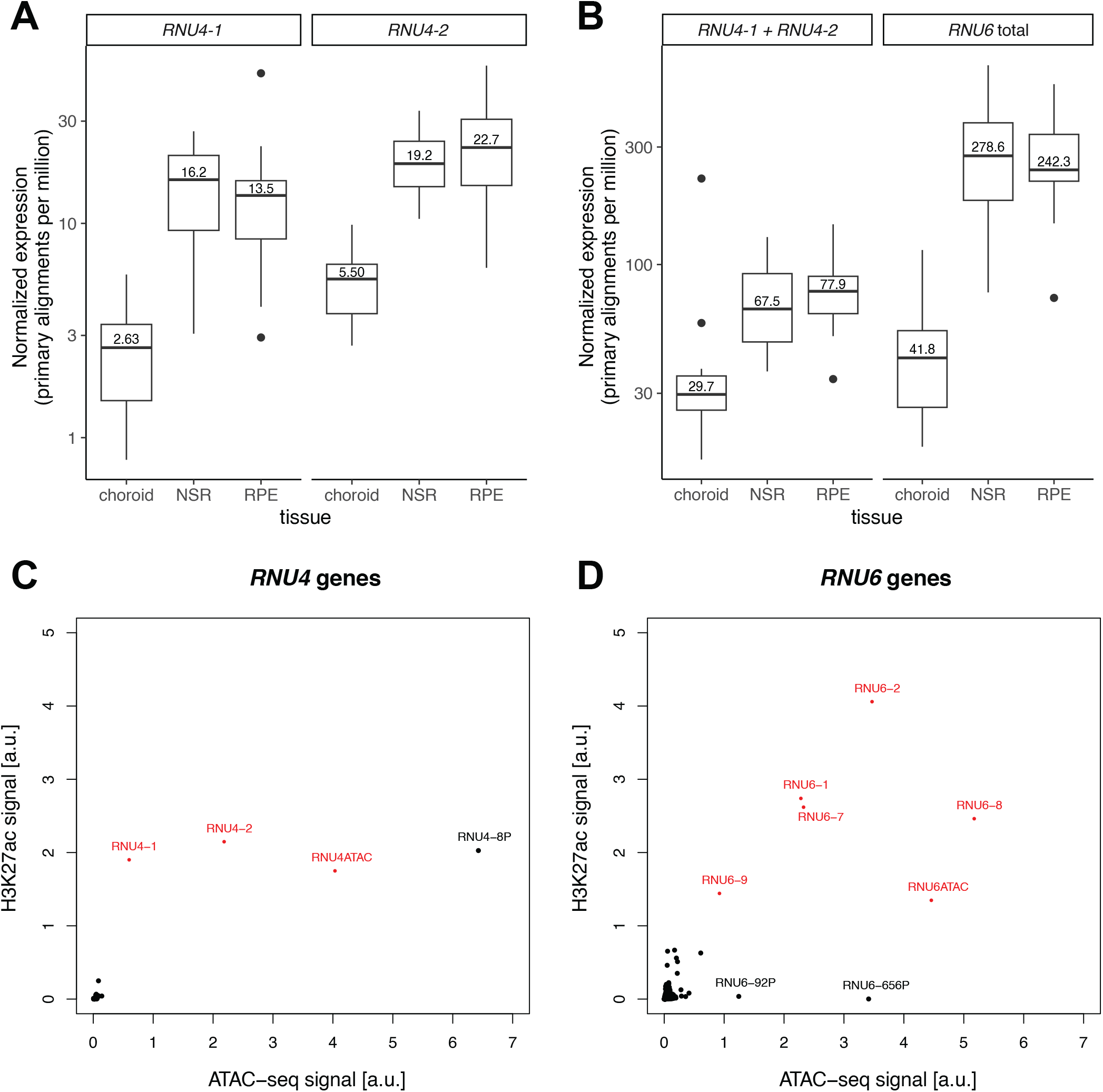
Expression and markers of transcriptional activity for *RNU4* and *RNU6* genes. (A) Expression of *RNU4-1* and *RNU4-2* from RNA-seq of human donor choroid (n=13), neurosensory retina (NSR, n=4) and retinal pigmented epithelium (RPE, n=16). Data are represented in boxplots and the median value is written in the box. (B) Same for *RNU4* genes (*RNU4-1, RNU4-2* and their pseudogenes (in black)) and for all *RNU6* genes (*RNU6-1, RNU6-2, RNU6-7, RNU6-8, RNU6-9* and their pseudogenes (in black)). (C) ATAC-seq and H3K27ac signals for three *RNU4* genes and 105 pseudogenes (D) ATAC-seq and H3K27ac signals for six *RNU6* genes and 1,312 pseudogenes. ATAC-seq data is from Wang *et al*.^54^ and H3K27ac data is from Cherry *et al*.^55^

In addition, we analyzed ATAC-seq and H3K27ac ChIP-seq data from retinal tissues^27^ in genomic regions spanning all *RNU4* and *RNU6* sequences. ATAC-seq assesses chromatin accessibility across the genome, while H3K27ac ChIP-seq reveals the presence of active enhancers. Combined, they provide indirect evidence of active transcription, even in the absence of reliable RNAseq data. Our analysis showed clear marks of active transcription in all paralogous genes (*RNU4-1, RNU4-2, RNU6-1, RNU6-2, RNU6-7, RNU6-8*, and *RNU6-9*) in the retina (Fig. 4C,D). Conversely, these signatures were absent from the 105 U4 pseudogenes and the 1,312 U6 pseudogenes, except for *RNU4-8P*, which displayed strong signals, but probably by virtue of its close proximity to the *ACTR1B* promoter. Of note, *RNU6-92P* and *RNU6-656P* had a high ATAC-seq signals but very low H3K27ac signals (Fig. 4C,D).

Likewise, we performed the same analysis for other snRNA genes present in the human genome, which revealed a similar trend: all *RNU* genes, with the exception of *RNU5F-1*, had marks of active transcription and only four (*RNU2-2P, RNU1-27P, RNU1-28P*, and *RNU5E-6P*) among the thousands of *RNU* pseudogenes displayed signals compatible with potential expression, therefore representing plausible candidate genes for retinal disease (Supplementary Fig. 4). Of note, *RNU2-2P* was recently associated with an NDD phenotype without a description of ocular features^12^. Interestingly, the same type of analysis, based on conservation and expression data from GTEx, was recently performed by others, showing similar results^28^.

For *RNU6-7*, both ATAC-seq and H3K27ac values were within the same range as those observed for other *RNU6* genes (Fig. 4D), and, therefore, the absence of pathogenic variants could not be explained by a potential differential expression. We thus analyzed the genetic landscape of variations in healthy individuals in all the five U6 paralogues, and observed that *RNU6-7* displayed a lower number of variants, compared to the others (Supplementary Fig. 5). In addition, we identified the recurrent variant n.55_56insG in *RNU6-7* in six control individuals of African / African American ancestry in gnomAD v4.1 (AF=0.014%) and in 14 individuals of African origin in the “All of Us” database (AF=0.013%). Considering these results, we propose that RNU6-7-derived U6 is either not part of the spliceosome or is present in only a small number of tri-snRNP complexes, a possibility that warrants investigation in future studies.

### Effect of RP variants on snRNP assembly

We immunopurified ectopically expressed U4 and U6 snRNAs containing the RP variants and analyzed their association with specific markers for U4/U6 di-snRNP (SART3 and PRPF31) and U5 snRNP (SNRNP200). All recurrent RP variants in U4 and U6 snRNAs increased the interaction between these snRNAs and di-snRNP markers (Fig. 5), while the U4 n.64_65insT change, which causes NDD, did not.

**Figure 5:**
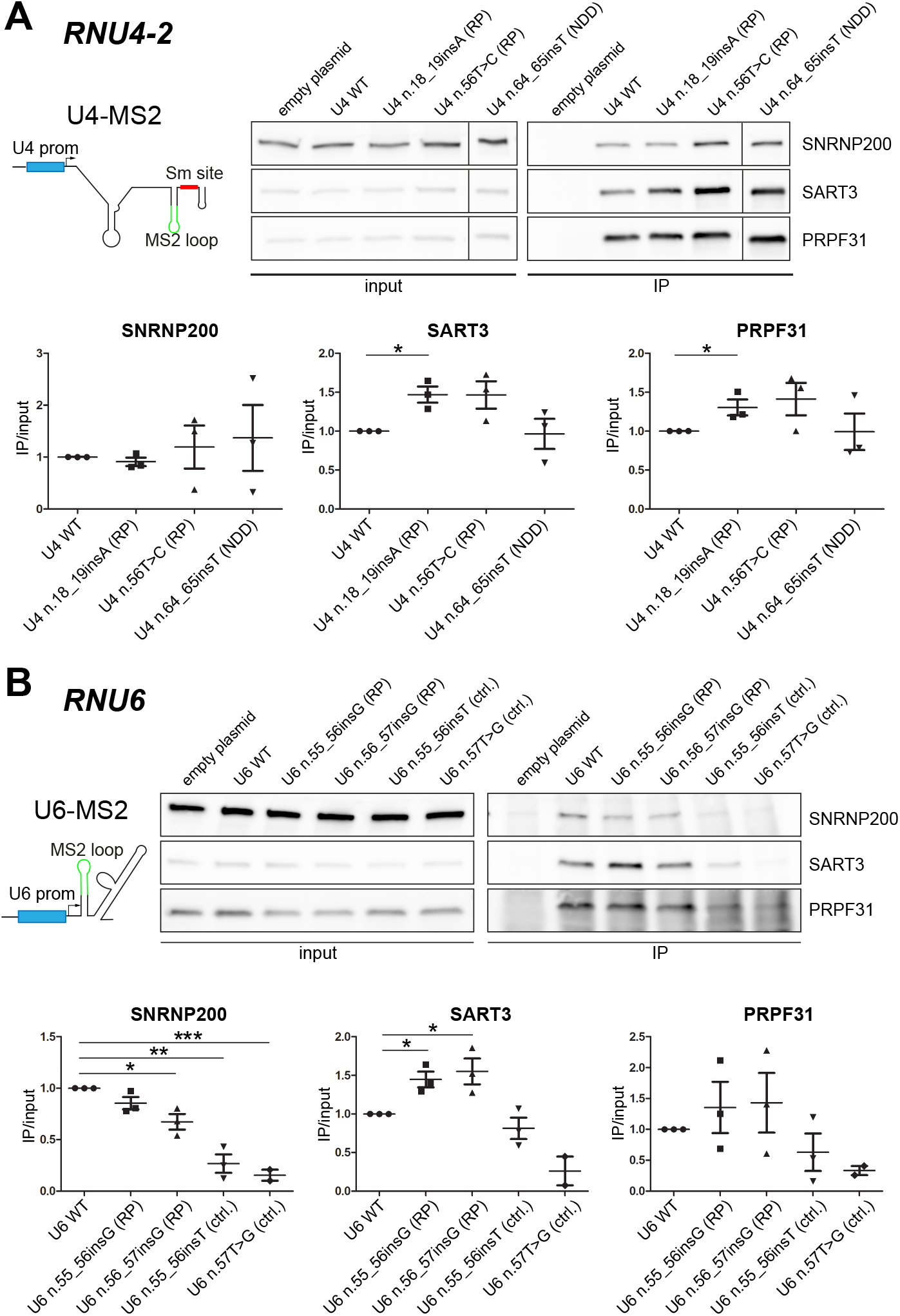
RP variants in *RNU4-2* and *RNU6* inhibit snRNP maturation. (A) Immunoprecipitation of U4-MS2 (WT and variants) and (B) U6-MS2 (WT and variants). snRNPs were immunoprecipitated via MS2-YFP by anti-GFP antibodies and co-precipitated proteins were detected by Western blotting. The position of the MS2 loop (green) in snRNAs is indicated. Three independent experiments were quantified. Immunoprecipitated proteins are normalized to input and U4 or U6 wild-type controls. Statistical significance was analyzed by the two-tailed unpaired t-test. * indicates P-value ≤0.05, ** ≤0.01, and *** ≤0.001

Similarly, U6 RNA bearing the n.55_56insT and n.57T>G variants, observed in healthy control individuals, did not affect the assembly process, and the low amount of protein associated with them indicates that they did enter the assembly process only minimally (Fig. 5B). Taken together, the results indicate that RP pathogenic variants have a specific dominant effect on snRNP biogenesis and slow down the assembly process at the di-snRNP stage.

## Discussion

The numerous genes associated with RP and allied diseases belong to very diverse functional classes, ranging from retina-restricted biochemical pathways to ubiquitous cellular processes. Although the molecular deficit resulting from specific RP variants can be modeled for many genes, the general mechanism of pathogenesis, linking thousands of DNA variants in dozens of genes to a common pathological phenotype, remains unknown, despite 40 years of intensive research. The correlation between pathogenic variants in splicing factors belonging to the tri-snRNP, essential for survival in all eukaryotes, and RP, a phenotype restricted to the eye, constitutes perhaps the most intriguing and complex of such molecular enigmas. Variants in these spliceosomal proteins (RP-PRPFs) are associated with the dominant form of the condition, the least prevalent one, at times with reduced penetrance^3,13^.

In this study, we have identified recurrent variants in *RNU4-2*, encoding U4 RNA, and in multiple paralogues of the U6 RNA, scattered across the whole human genome, as a cause of RP. Interestingly, these snRNAs are also an integral part of the di- and tri-snRNP and directly interact with some RP-PRPF proteins. In addition, like RP-PRPFs, they are also associated with the same specific phenotype: *de novo* or inherited adRP, with reduced penetrance for *RNU4-2* variants. The clinical presentation of patients with *RNU4-2* and *RNU6* variants from this study exhibit as well overlap with the other known forms of spliceosome-related adRP, particularly in terms of earlier disease onset (contrasting the generally milder prognosis of most other forms of adRP^29,30^) and the relatively high co-occurrence of features such as cataracts and CME found in *PRPF31*^*31,32*^, *PRPF8*^*33*^ and *SNRNP200*^*34*^. Prevalence estimations indicate that these snRNA pathogenic changes may account for an elevated number of undiagnosed cases, and it is therefore surprising that the *RNU4* and *RNU6* genes have escaped disease association until now. A partial explanation for this phenomenon is that mainstream sequencing approaches, both in research and diagnostics, are biased towards DNA-capturing procedures that do not include snRNA genes. Nevertheless, genome sequencing is being implemented in routine diagnostics, and these variants may have eluded detection so far because they affect non-coding transcripts and are therefore more difficult to score in terms of potential pathogenicity by analytical pipelines.

An intriguing feature of pathogenic changes in *RNU4-2* is their pleiotropy with respect to NDD (ReNU) and RP. Although the exact mechanism for this phenotypic selectivity is unknown, it highlights the presence of a new allelic series involving non-coding RNA genes. A potential explanation for such divergent phenotypes could reside in the position that ReNU and RP variants have with respect to the U4/U6 duplex. ReNU pathogenic variants are included within the stem III of the duplex, whereas RP variants cluster, in close spatial proximity, to the three-way junction, in regions to which several of the RP-associated splicing factors bind. Our ectopic expression experiments showed in fact that snRNAs bearing RP variants display enhanced interaction with di-snRNP protein markers, suggesting that pathogenesis could result from a gain-of-function / dominant-negative mechanism, rather than from haploinsufficiency. This hypothesis was strengthened by the evidence that molecularly similar but benign variants, commonly observed in the general population, seemed not to efficiently bind to di-snRNP markers, supporting the idea that spliceosomal functions could be haplosufficient with respect to heterozygous and snRNA-depleting variants. Moreover, the pathogenic variants identified in this study are located precisely in regions of the U4/U6 duplex that interact directly with PRPF3 and PRPF31 proteins, two splicing factors previously associated with adRP. These proteins, when mutated, destabilize the spliceosome complex formation, leading to delayed and abnormal splicing events, particularly in the retina^26,35,36^. Therefore, it is plausible that all RP variants identified in *RNU4-2* and *RNU6* paralogues could lead to photoreceptor death and subsequent visual loss via similar early events of pathogenesis, while having no influence on the development of the brain. The fact that RP variants in *RNU4-2* are not located in the variant-depleted region of the gene that defines the NDD hotspot also suggests that such variants may be milder with respect to those giving rise to ReNU, leading to delays in complex assembly rather than to defects in pre-mRNA processing.

Although variants associating *RNU4-2* to ReNU syndrome have been primarily reported as *de novo* events^8,12^, in our study most individuals with RP (67%) bear *RNU4-2* and *RNU6* changes as inherited variants. In part, this difference can be explained by the reduced reproductive fitness associated with NDD. Unlike ReNU syndrome, symptomatic onset (night-blindness and peripheral vision loss) in non-syndromic adRP begins later in life, with severe central vision loss occurring significantly beyond reproductive age. Another difference involves the inheritance of dominant variants, which in ReNU seems to be almost exclusively of maternal origin^8^. We did not observe the same trend for RP, with variants being inherited from fathers and mothers in numerous cases, indicating the absence of any sex-specific negative selection during gametogenesis or at the embryonic stage.

The human genome contains two *RNU4* paralogues and five *RNU6* paralogues. This indicates that, assuming equal expression within paralogues, the presence of only ∼25% of mutant U4 (heterozygous genotype, over two copies) or ∼10% of mutant U6 (heterozygous genotype, over five copies) is sufficient to lead to a disease phenotype, again in support of a gain-of-function / dominant-negative molecular mechanism. This could be a crucial consideration for the development of potential gene-based therapies, as gene-augmentation strategies may be suboptimal compared to gene correction or antisense oligonucleotide approaches. Our data also highlight the existence of mutational hotspots outside the coding regions of the human genome, emphasizing the need for further research into these parts of our genetic material, and show that the clustering of *de novo* pathogenic variants is not restricted to severe diseases with childhood onset^37^, but may extend to milder pathologies, such as RP.

In conclusion, we identified four recurrent pathogenic variants in *RNU4-2* and four out of the five paralogues of the U6 snRNA as a frequent cause of *de novo* or inherited adRP. The immediate impact of these findings involves improved diagnosis and genetic counselling for patients with hereditary visual loss, especially for isolated cases who could potentially bear heterozygous *de novo* events. More fundamentally, this work significantly broadens our understanding of the genetic landscape of human disease, paving the way for the development of new molecular therapeutic approaches.

## Supplementary Methods

### Patients and DNAs

This study adhered to the tenets of the Declaration of Helsinki and complied with the ethical standards of each participating country. Signed informed consent forms were obtained in all study subjects. Our research was approved by the Ethikkommission Nordwest-und Zentralschweiz (Project-ID 2019-01660) and the Ethics Committee of the Radboud University Medical Center Nijmegen (MEC-2010-359; OZR protocol no. 2009-32).

### Clinical characterization and analysis

Complete ophthalmic examinations were performed by a retinal specialist, which included measurement of best-corrected visual acuity (BCVA), intraocular pressures (IOP), and examination of anterior segment and fundus (dilated). Color fundus photographs and montages were captured using the FF450plus Fundus Camera (Carl Zeiss Meditec, Germany) and Optos 200 Tx (Optos PLC, UK). Fundus autofluorescence FAF images (488-nm excitation) and high-resolution spectral-domain optical coherence tomography (SD-OCT) scans were acquired using the Spectralis HRA+OCT module (Heidelberg Engineering, Heidelberg, Germany). Hyper-autofluorescent ring contour were analyzed using a custom program in FIJI software (National Institute of Mental Health, Bethesda, MD, USA) as previously described^38^. Progression rates were calculated using linear mixed-effects regression in R (version 4.0.4) with time (years) since baseline as the primary independent variable, baseline ring size as a covariate and inter-ocular differences as a random effect. Photoreceptor+ thickness was assessed on horizontal SD-OCT scans through the fovea using a semiautomated procedure previously described^39^. Photoreceptor+ was defined as the distance between the Bruch’s membrane/choroid interface and the inner nuclear layer (INL)/outer plexiform layer (OPL) boundary. Layer segmentation was performed in a semi-automated manner using a custom software in MatLab (MathWorks, Inc). Full-field electroretinogram recordings were conducted using the Espion Visual Electrophysiology System (Diagnosys LLC) according to International Society for Clinical Electrophysiology of Vision (ISCEV) standards^40^.

### Genome sequencing and annotation

Genomic DNA from probands was isolated from peripheral blood lymphocytes according to standard procedures. Sequencing was performed by BGI Tech Solutions (Warszawa, Poland) using the DNBseq Sequencing Technology, with a minimal median coverage per genome of 30x. The processing of the sequencing data (mapping, variant calling, and variant annotation) was performed as described previously^2^. Briefly, we used BWA^41^, Picard (http://broadinstitute.github.io/picard) and GATK^42^ for mapping and variant calling. For variant annotation, we used ANNOVAR^43^ with the addition of splicing predictions by MaxEntScan^44^ and SpliceAI^45^.

### Assessment of variants

HGVS notations of the variants were retrieved using VariantValidator^46^ and ACMG classification^47^ was applied according to the ACGS Best Practice Guidelines for Variant Classification in Rare Disease 2023^48^. All pathogenic and likely pathogenic variants are listed in Supplementary Table 1, all VUSs and likely benign variants that were reported are listed in Supplementary Table 4.

### Screening by Sanger sequencing

Genomic DNA was collected, and *RNU4-1, RNU4-2, RNU6-1, RNU6-2, RNU6-7, RNU6-8* and *RNU6-9* genes were amplified using standard PCR procedures. *RNU4-1, RNU4-2, RNU6-1, RNU6-2, RNU6-7, RNU6-8* and *RNU6-9* PCR fragments were sequenced using Sanger sequencing and screened for novel variants in these genes. Sequences of primers used in this study are listed in the Supplementary Table 6. Additional details regarding PCR conditions or primer design are available upon request.

### 2D modeling of the effect of variants and 3D representation

We utilized RNAfold WebServer to model the effect of variants with default parameters^49^ and RNAcanvas was used for drawing the structure^50^. We used ChimeraX with PDB file using PDB file 6qw6 to draw 3D representation of the U4/U6 duplex with and without surrounding PRPF proteins.

### RNA-seq experiments and analysis

RNA was isolated from human donor eye tissue, which was collected and dissected as described previously^51^ from an ethically approved Research Tissue Bank (UK NHS Health Research Authority reference no. 15/NW/0932). Total RNA was isolated from four neurosensory retina (NSR) samples, 16 pelleted retinal pigment epithelium (RPE) samples and 13 choroid samples that had been stored in RNAlater (Thermo Fisher Scientific, Carlsbad, CA, USA), using an Animal Tissue RNA Purification kit (Norgen Biotek, Canada), as per manufacturer’s instructions. Sequencing libraries were prepared using the NEBnext multiplex small RNA library preparation kit, as per manufacturer’s protocols, with size selection performed using Ampure beads. Paired-end sequencing (2×75bp) was performed on an Illumina HiSeq 4000.

NEBnext adapters were removed from sequencing reads using trimmomatic prior to alignment against the GRCh38 reference genome with bowtie^52^. No mismatches between sequencing reads and the reference genome were allowed, and no restriction was set on multi-mapping reads. Sequence read counts were restricted to primary alignments using samtools v1.9^53^, and therefore only counted once if they aligned to multiple *RNU4* or *RNU6* genes or pseudogenes. Calculations were drawn from read1 datasets, and normalised for the total read count achieved for the sample. Total RNU4 and RNU6 expression was based on all annotated genes and pseudogenes in gencode.

### ATAC-seq and H3K27ac data

ATAC-seq data from Wang *et al*.^*54*^ (eight different experiments) and H3K27ac data from Cherry *et al*.^*55*^ (five different experiments) were downloaded as bigwig files from the RegRet database (http://genome.ucsc.edu/s/stvdsomp/RegRet)^56^. For both data types, the signal (the genes and 500bp on each side) was extracted using bedtools after conversion using bigWigToWig. We quantified the signal for all RNU genes and pseudogenes first by normalizing the signal of each experiment to the maximum and then summing them. For RNU4, we quantified 3 genes and 105 pseudogenes while for RNU6, it was 6 genes and 1,312 pseudogenes.

### U4 and U6 snRNP analysis

U4 n.18_19insA, n.56T>C, and n.64_65insT variants were introduced by site-directed mutagenesis into the plasmid expressing U4-MS2 described previously^57^. The full-length U6 sequence, including 256 bp upstream and 93 bp downstream of the *RNU6-1* gene, was inserted into the pcDNA3 plasmid lacking the CMV promoter. The MS2 loop was inserted between nucleotides 10 and 11. U6 n.55_56insG, n.55_56insT, n.56_57insG, and n.57G>T variants were introduced by site-directed mutagenesis. U4- and U6-expressing plasmids were transfected into HeLa cells stably expressing MS2-YFP protein. 24 hours after transfection, snRNAs were immunoprecipitated using anti-GFP antibodies and co-precipitated proteins were analyzed by Western blotting as described previously^57^.

## Data availability

Research on the de-identified patient data used in this publication from the Genomics England 100,000 Genomes Project and the NHS GMS dataset can be carried out in the Genomics England Research Environment subject to a collaborative agreement that adheres to patient-led governance. All interested readers will be able to access the data in the same manner that the authors accessed the data. For more information about accessing the data, interested readers may contact or access the relevant information on the Genomics England website: https://www.genomicsengland.co.uk/research. The data generated during this study (pathogenic variants and VUSs identified) are submitted to the Leiden Open (source) Variation Database (LOVD) (http://www.lovd.nl) and ClinVar.

## Acknowledgements and funding sources

The authors of the article would like to thank the patients and their family members for their help and participation in this study. We are very grateful to Sitta Föhr for technical and administrative support, to Saskia van de Velde-Visser, Ellen Blokland and Marlie Jacobs-Camps for sample registration and administration, to Mattias Van Heetvelde, Toon Rosseel (computational), Quinten Mahieu (technical), Marieke Debruyne, Stijn Van de Sompele (clinical reporting) for their support.

C.R. was supported by the Swiss National Science Foundation (SNSF) Grant No. 310030_204285 entitled “Genomics of inherited retinal diseases”. S.R. was supported by the Foundation Fighting Blindness Career Development Award (CD-GE-0621-0809-RAD), Radboudumc Starter grant (OZI-23.009) and NWO Aspasia (015.021.028). S.R., C.R., E.D.B., M.B., S.B., D.St., were supported by HORIZON-MSCA-2022-DN (No.101120562, ProgRET). E.D.B., S.R., C.R. were supported by the EJPRD19-234 Solve-RET. This work has been funded by a Foundation Fighting Blindness Program Project Award (PPA-0622–0841-UCL) (to A.J.H, S.R. and S.E.dB.). S.R. and F.P.M.C. were supported by the Gelderse Blindenstichting, the Algemene Nederlandse Vereniging ter voorkoming van Blindheid, Oogfonds, Landelijke Stichting voor Blinden en Slechtzienden, Rotterdamse Stichting Blindenbelangen, Stichting Blindenhulp, Stichting tot Verbetering van het Lot der Blinden, and Stichting Blinden-Penning. M.Q. was supported by the RetinAward 2021. S.H.T. - Jonas Children’s Vision Care (JCVC) is supported by the National Institute of Health U01EY030580, U01EY034590 R24EY028758, R24EY027285, 5P30EY019007, R01EY033770, R01EY018213, R01EY024698, the Foundation Fighting Blindness TA-GT-0321-0802-COLU-TRAP, Richard Jaffe, NYEE Foundation, the Rosenbaum Family Foundation, the Gebroe Family Foundation, Piyada Phanaphat Fund, the Research to Prevent Blindness (RPB) Physician-Scientist Award, unrestricted funds from RPB, New York, NY, USA. C.A. was supported by Instituto de Salud Carlos III (ISCIII) of the Ministerio de Ciencia e Innovación and Unión Europea – European Regional Development Fund (FEDER) (PI22/00321 and IMP/00009), Centro de Investigación Biomédica en Red Enfermedades Raras (CIBERER, 06/07/0036), IIS-FJD BioBank (PT13/0010/0012), the Organización Nacional de Ciegos Españoles (ONCE), European Regional Development Fund (FEDER), the University Chair UAM-IIS-FJD of Genomic Medicine. This work was performed by using the data contained in the “Programa Infraestructura de Medicina de Precisión asociada a la Ciencia y la Tecnología en Medicina Genómica (IMPaCT-GENóMICA)”, coordinated by the CIBERER and founded by ISCIII. L.F.C. was supported by Centro de Investigación Biomédica en Red (CIBER). R.A. was supported by the National Eye Institute (NEI) (RO1 EY030591, RO1 EY031663, T32 EY026590, and P30 EY22589). C.C.W.K was supported by the Combined Ophthalmic Research Rotterdam) grant 8.2.0. S.B. was supported by the Italian Telethon Foundation. G.J.F. and N.C. were supported by Fighting Blindness Ireland (FB22FAR, FB16FAR), Fighting Blindness Ireland - Health Research Charities Ireland (MRCG-2016-14) and Science Foundation Ireland (16/IA/4452 and 22/FFP-A/10544). J.E. was supported by the Macular Society (United Kingdom), and the University of Manchester Core Genomics Technology Facility. T.B.Y. was supported by the Israel Science Foundation (331/24). A.C.B.J. is supported by the University of Melbourne Research Fellowship. K.M.B. was supported by the National Eye Institute (NEI) (RO1 EY035717 and P30 EY014104 [MEE core support]), the Iraty Award 2023, the Lions Foundation, and the Research to Prevent Blindness (Unrestricted Grant). L.S.S., E.L.C. and S.P.D. were supported by grants from the Foundation Fighting Blindness (EGI-GE1218-0753-UCSD) and the Brett & Jane Eberle Foundation. E.D.B. and B.P.L were supported by Ghent University Special Research Fund (BOF20/GOA/023) and E.D.B. (1802220N) and B.P.L. (1803816N) are Senior Clinical Investigators of the Research Foundation-Flanders (FWO). N.M. and S.S. are Ph.D. fellows of HORIZON-MSCA-2022-DN ProgRET (No.101120562). R.A. was supported by the Foundation Fighting Blindness. J.D was supported by the UCSF Vision Core shared resource of the NIH/NEI P30 EY002162, the Foundation Fighting Blindness, and an unrestricted grant from Research to Prevent Blindness. T.I. was supported by research grant from the Japan Agency for Medical Research and Development (AMED) (20ek0109493h0001, 21ek0109493h0002, 22ek0109493h0003, 23ek0109617h0002, 24ek0109617h0003). R.K.K. was supported by The Montreal Children’s Hospital Foundation, The Vision Sciences Research Network (VSRN), The National Institutes of Health (NIH)(R01 EY030499-01, Dr. Lentz), The Canadian Institutes for Health Research (CIHR), Fighting Blindness Canada (FBC), and Fonds de Recherche du Québec – Santé (FRQS). R.K.K. participates in the NAC Attack clinical trial, which is funded by the National Institutes of Health via grants UG1EY033286, UG1EY033293, UG1EY033286, and UG1EY033292. T.M.L., T.L.M and J.N.D.R. were supported by Retina Australia (awarded to the Australian Inherited Retinal Disease Registry and DNA Bank). O.M. was supported by the Wellcome Trust (grant no: 206619/Z/17/Z). T.L.M. was supported by Retina Australia. Awarded to the Australian Inherited Retinal Disease Registry and DNA Bank. M.P. was supported by the BrightFocus Foundation (M2024009N). E.A.P. was supported by the National Eye Institute (NEI) (R01 EY012910). R.R. was supported by Retina South Africa and the South African Medical Research Council (MRC). S.G.S. and E.M.V. were supported by the Italian Ministry of Health (RF-2019-12369368). D.St. was supported by the Ministry of Education, Youth and Sports of the Czech Republic grant for RNA therapy (CZ.02.01.01/00/22_008/0004575). T.B.H. was supported by the European Commission (Recon4IMD - GAP-101080997) and the Deutsche Forschungsgemeinschaft (German Research Foundation, DFG, grant numbers 418081722 and 433158657 to T.B.H.). P.L. and L.D. were supported by a research grant (NW24-06-00083) from the Ministry of Health of the Czech Republic and UNCE/24/MED/022. V.R.dJ.L.R. and J.C.Z. were supported by the Velux Stiftung Grant 1860. M.A., M.M., C.F.I. J.C.G, A.J.H, and C.T. are supported by Retina UK and Fight for Sight UK (RP Genome Project Grant GR586). A.J.H, J.C.G, M.M, O.A.M, A.W, G.A and S.L were supported by the National Institute for Health Research Biomedical Research Centre at Moorfields Eye Hospital and UCL Institute of Ophthalmology. S.L. was funded by an MRC Clinician Scientist Fellowship. J.M.S. was supported by Instituto de Salud Carlos III (ISCIII) of the Spanish Ministry of Health (PI22/00213), Centro de Investigación Biomédica en Red Enfermedades Raras (CIBERER, 06/07/1030, CIPROM/2023/26 from the Generalitat Valenciana and IMPaCT-GENOMICA (IMP/00009) co-funded by ISCIII and FEDER. S.R., C.C.W.K, C.J.F.B., A.G. were supported by the Dutch ministry of Education, Culture and Sciences, Gravitation grant: 024.006.034 Lifelong VISION. W.L. was supported by the National Institute of Health/National Eye Institute (1K99EY036930-01).

This work is supported by partners of the European Reference Network for Rare Eye Diseases ERN-EYE (Grant Agreement No 101085439, C.J.F.B., E.D.B., C.B.H, S.K, B.P.L, P.L., L.H-W., K.S, L.I.v.d.B.). Novartis contributed funding for the preceding RP-LCA smMIPs panel design and subsequent sequencing (to F.P.M.C., S.R. and D.M.P). Novartis was not involved in the study design, collection, analysis, interpretation of data, the writing of this article, or the decision to submit it for publication.

This research was made possible through access to data in the National Genomic Research Library, which is managed by Genomics England Limited (a wholly owned company of the Department of Health and Social Care). The National Genomic Research Library holds data provided by patients and collected by the NHS as part of their care, and data collected as part of their participation in research. The National Genomic Research Library is funded by the National Institute for Health Research and NHS England. The Wellcome Trust, Cancer Research UK, and the Medical Research Council have also funded research infrastructure.

