## Supplementary material for "De novo and inherited dominant variants in U4 and U6 snRNAs cause retinitis pigmentosa": Figure S1

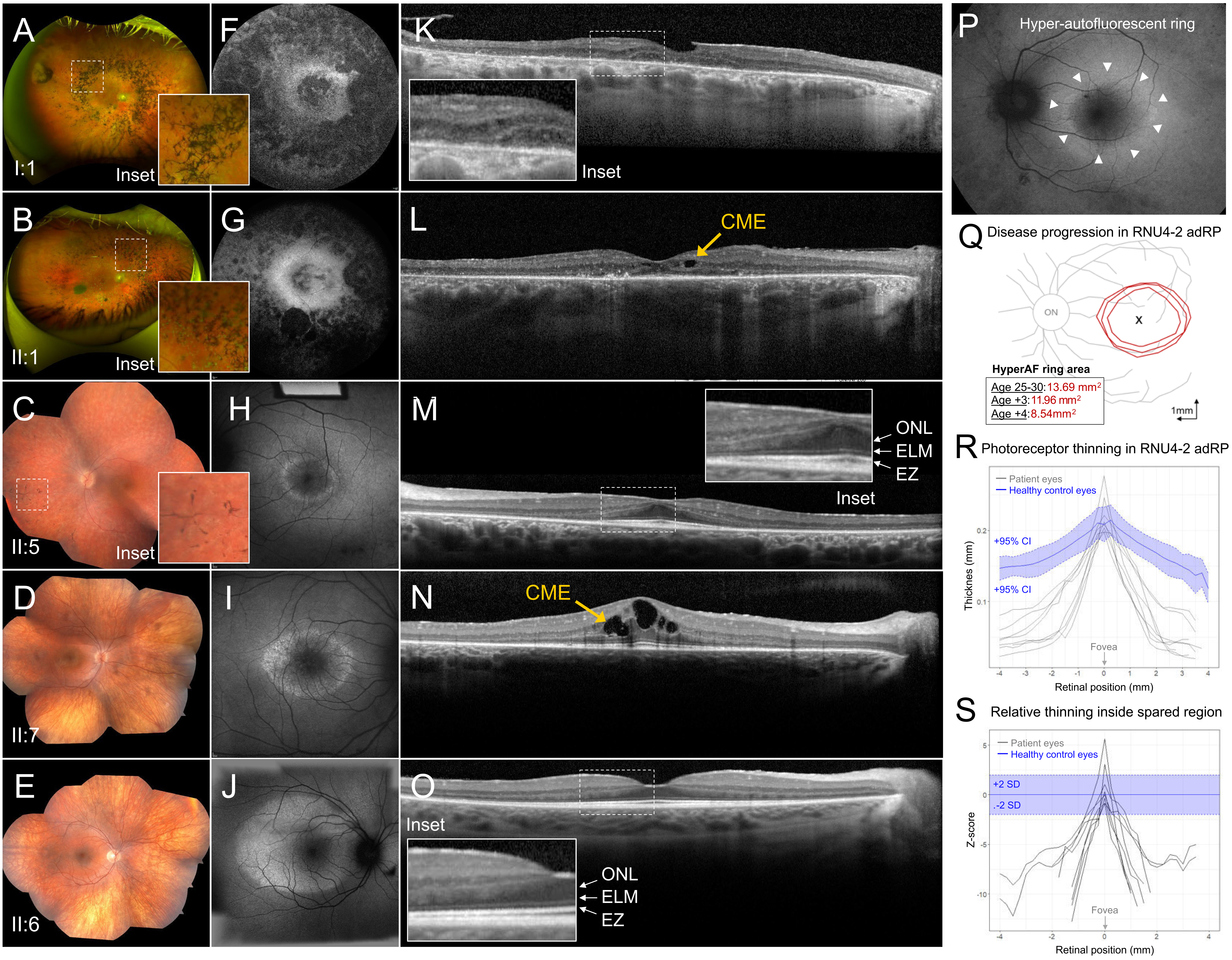


**Supplementary Figure 1: Clinical images of the initial family with adRP with an *RNU4-2* variant.** Clinical phenotype of the affected father (I:1) and representative affected siblings, I:1, II:5, II:7 and II:6 in the original family harboring the pathogenic variant n.18_19insA in *RNU4-2*. (A-E) Wide-field and color fundus montage photographs show varying stages of degeneration and bone-spicule pigment deposition (inset) across the midperiphery. Fundus autofluorescence (FAF) showing foveal atrophy (F-G) and the characteristic autofluorescence ring (H-J) delineating area of spared retina. (K-O) Horizontal spectral-domain optical coherence tomography (SD-OCT) of structurally intact outer retinal layers and the presence of cystoid macular edema (L, N). (P) FAF image delineating the contour of the autofluorescent ring (white arrowheads) in II:2. (Q) Contour diagram tracing progressive constriction of the autofluorescent ring over a 4 year period in II:6 (ON, optic nerve; x, fovea). (R) Thickness profiles of photoreceptor+ on SD-OCT scans across the macula in siblings with visible autofluorescent rings (II:6, II:2, II:3 and II:7) (gray lines) compared to the mean and 95% confidence intervals (CI) (shaded region) of 20 age-matched healthy control eyes. (S) Normalized photoreceptor+ thicknesses within the autofluorescent ring relative to ± 2 standard deviations (SD) of healthy control eyes.
