## Supplementary material for "De novo and inherited dominant variants in U4 and U6 snRNAs cause retinitis pigmentosa": Figure S2

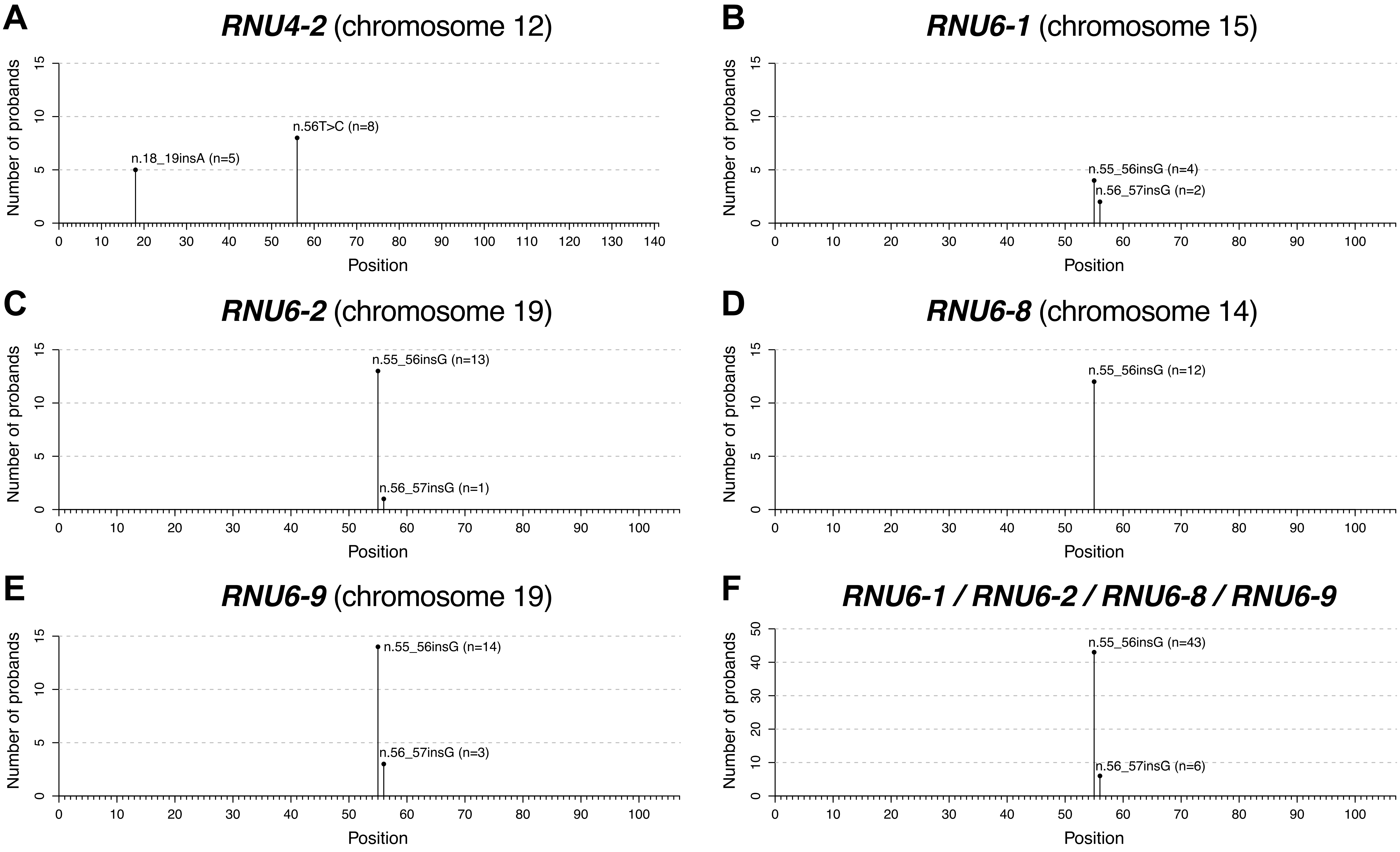


**Supplementary Figure 2: Recurrent pathogenic variants identified in RP cases** in (A) *RNU4-2,* (B) *RNU6-1,* (C) *RNU6-2,* (D) *RNU6-8* and (E) *RNU6-9* observed in RP cases. (F) Total for *RNU6-1, RNU6-2, RNU6-8* and *RNU6-9*.
