## Supplementary material for "De novo and inherited dominant variants in U4 and U6 snRNAs cause retinitis pigmentosa": Figure S3

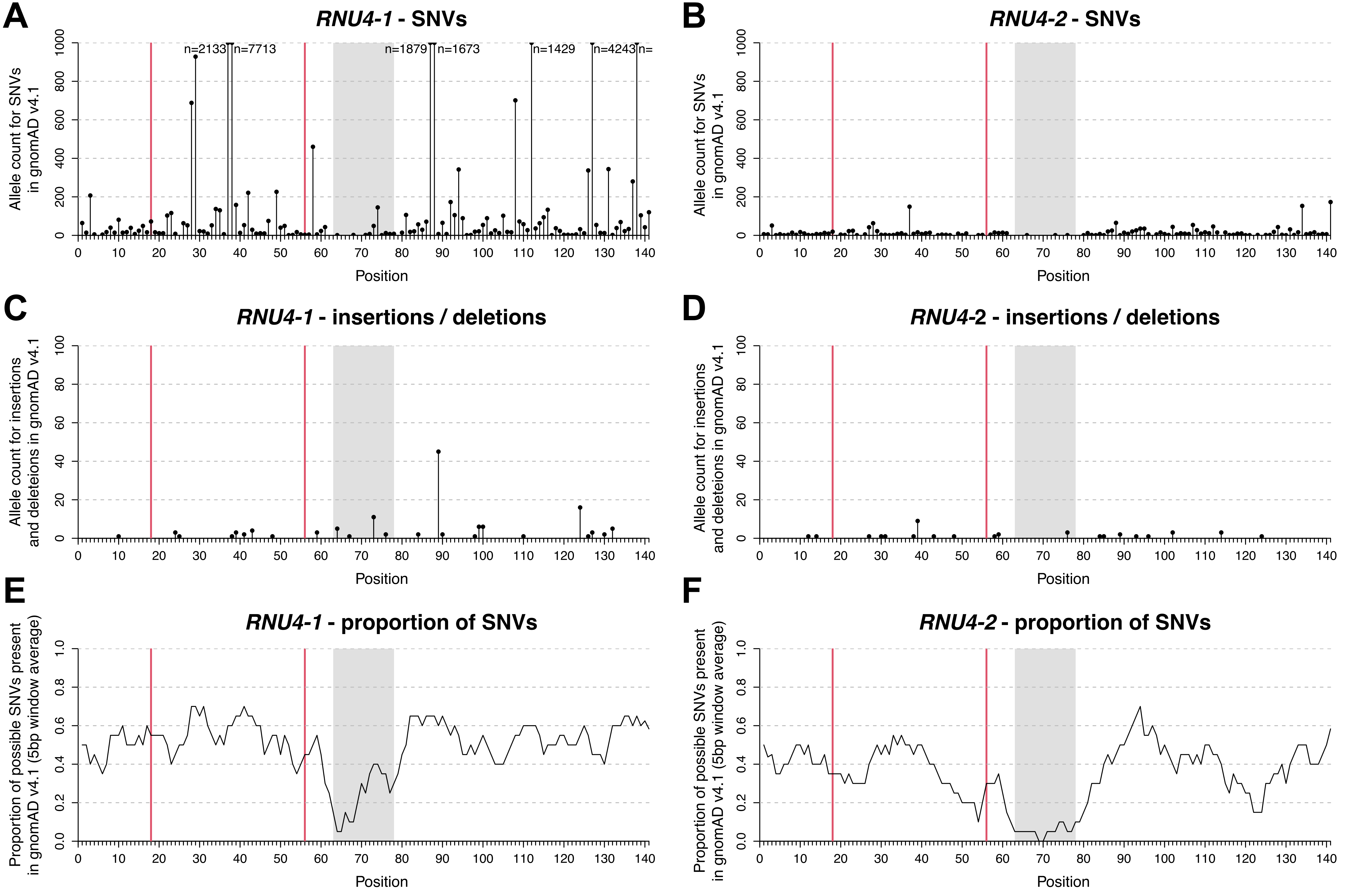


**Supplementary Figure 3: Landscape of variations from gnomAD v4.1 for *RNU4-1* and *RNU4-2*.** (A, B) Allele count of SNVs for *RNU4-1* and *RNU4-2*. (C, D) Allele count of insertions and deletions for *RNU4-1* and *RNU4-2*. (E, F) Proportion of possible SNVs for *RNU4-1* and *RNU4-2* with a 5bp running window average. The position of recurrent pathogenic variants causing RP detected in *RNU4-2* are indicated with red lines. The region that contains pathogenic variants in *RNU4-2* causing NDD is shaded in grey.
