## Supplementary material for "De novo and inherited dominant variants in U4 and U6 snRNAs cause retinitis pigmentosa": Figure S4

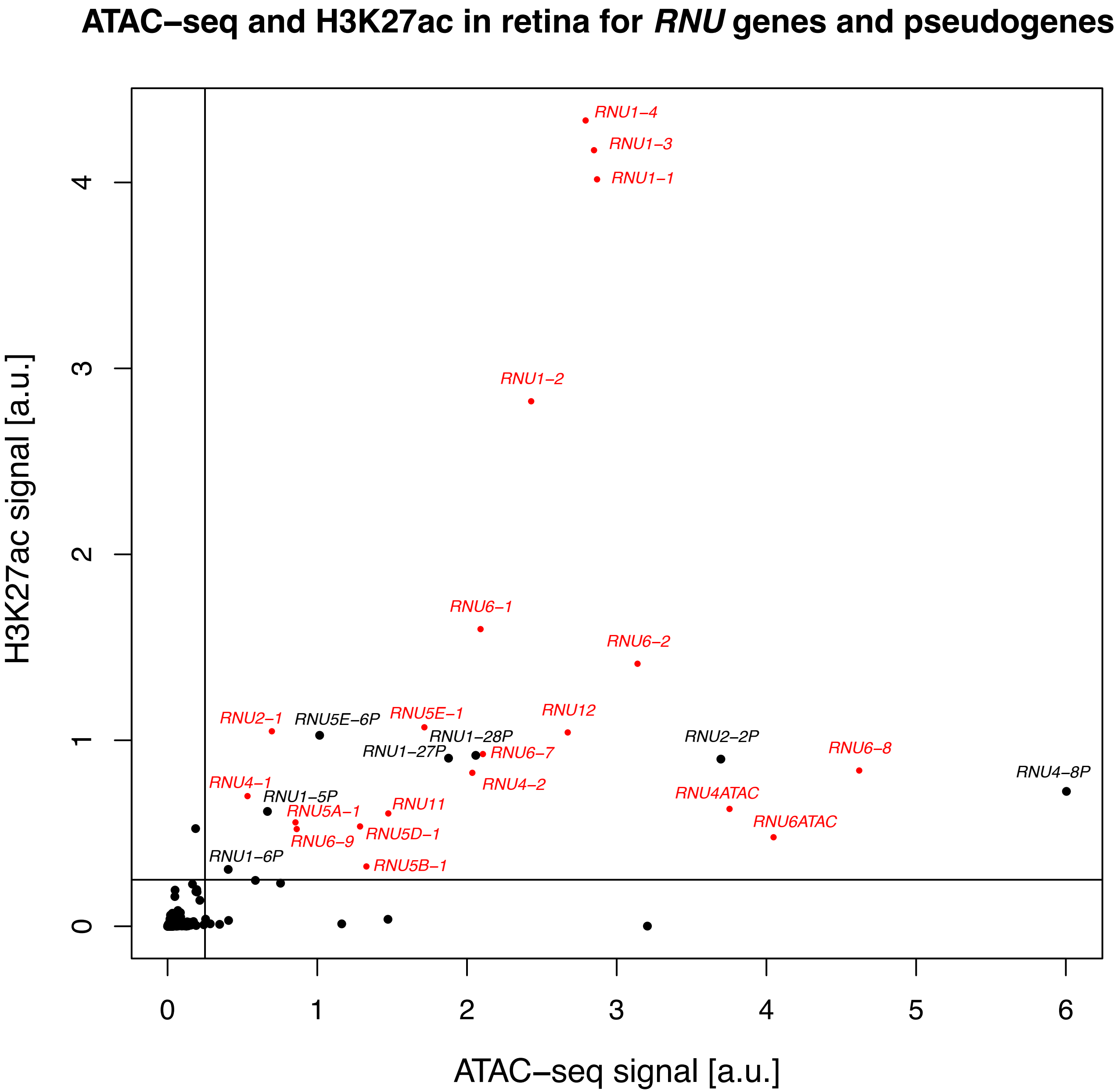


**Supplementary Figure 4: Markers of transcriptional activity in human retinal tissue in the regions of all *RNU* genes and pseudogenes.** Gene (in black) and pseudogene (in red) names are mentioned when both signals are higher than 0.25 (vertical and horizontal black lines). ATAC-seq signal from Wang *et al.*[54] and H3K27ac data is derived from Cherry *et al.*[55]
