## Supplementary material for "De novo and inherited dominant variants in U4 and U6 snRNAs cause retinitis pigmentosa": Figure S5

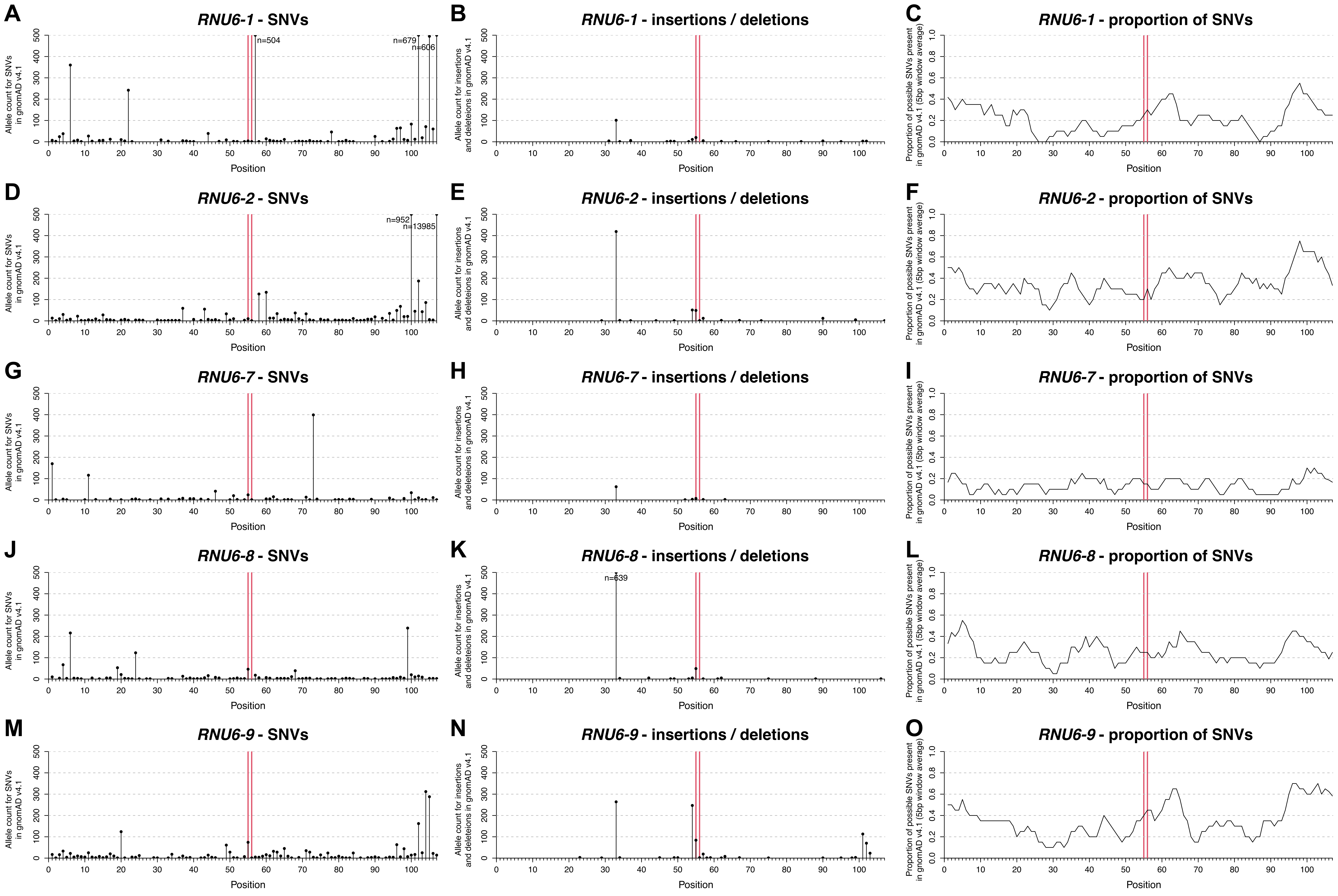


**Supplementary Figure 5: Landscape of variations from gnomAD v4.1 for *RNU6-1, RNU6-2, RNU6-7, RNU6-8,* and *RNU6-9*.** (A, D, G, J, M) Allele count of SNVs of the different genes. (B, E, H, K, N) Allele count of insertions and deletions. (C, F, I, L, O) Proportion of possible SNVs with a 5bp running window average. The position of recurrent pathogenic variants underlying RP identified are indicated with red lines.
