## Supplementary Data 1 for "De novo and inherited dominant variants in U4 and U6 snRNAs cause retinitis pigmentosa"

We initially examined a non-consanguineous family with autosomal dominant RP spanning two generations. Seven of eight siblings (II:1-II:7) and their father (I:1) were affected, each experiencing a symptomatic onset of night-blindness and progressive loss of peripheral vision beginning in late adolescence to early adulthood. Posterior subcapsular cataracts developed in 5 siblings (II:1-II:5) and I:1 by their 3rd decade. Fundus examination in all affected individuals revealed classical RP features, including waxy disc pallor, attenuated retinal vessels and bone-spicule pigment (BSP) deposition. BSP distribution was most pronounced in the superonasal quadrant, varying in severity from mild specks in the midperiphery to diffuse migration across the posterior pole (Supplementary Fig. 1A-E). Best-corrected visual acuities (BCVA) were stable from 20/20 to 20/60 in siblings, except I:1, whose BCVA declined to hand motion/light perception between the 6th and 7th decades. During the follow up period, I:1, II:1, II:2, II:4 and II:7 developed persistent cystoid macular edema (CME) and further contributed to central vision loss. Fundus autofluorescence (FAF) imaging showed widespread coalescing lesions of RPE atrophy in the periphery with progressive involvement of the fovea in I:1 and II:1 (Supplementary Fig. 1F-G); the spared region of residual function in siblings II:2-II:7 was delineated by a characteristic hyper-autofluorescent “ring” that ranged in size from 5.32 to 50.17 mm^2^ and constricted at a rate of 0.48 to 1.25 mm^2^/year (Supplementary Fig. 1H-J, P, Q) inside the macula. Both RPE and photoreceptor-attributable layers [cone outer segment tips (COST), ellipsoid zone (EZ), external limiting membrane (ELM) and outer nuclear layer (ONL)] were structurally intact within this spared region (Supplementary Fig. 1K-O), although significantly thinned relative to spatially corresponding regions of a healthy retina (Supplementary Fig. 1R-S). In all individuals, dark-adapted (scotopic) 0.01 cd·s/m² responses were extinguished on full-field electroretinogram (ffERG) testing indicating generalized rod dysfunction. Mixed rod and cone dark-adapted 3.0 cd·s/m² scotopic waveforms were recordable but severely attenuated (b-wave~17 to 80μV) in all except I:1 (extinguished). Light-adapted (photopic) responses were comparatively preserved but nevertheless affected in all cases indicating subsequent cone involvement. 30-Hz flicker peak-to-trough amplitudes ranged from 10 to 50 μV and demonstrated delayed implicit time of >40 ms.
